# Associations of infections and vaccines with Alzheimer’s disease point to a major role of compromised immunity rather than specific pathogen in AD

**DOI:** 10.1101/2023.12.04.23299092

**Authors:** Svetlana Ukraintseva, Arseniy P. Yashkin, Igor Akushevich, Konstantin Arbeev, Hongzhe Duan, Galina Gorbunova, Eric Stallard, Anatoliy Yashin

**Author notes:** **To whom correspondence should be addressed:** Dr. Svetlana Ukraintseva, Biodemography of Aging Research Unit (BARU), Duke University, PO Box 90420, 2024 W. Main St., Durham, NC 27708, Dr. Arseniy P. Yashkin, Biodemography of Aging Research Unit (BARU), Duke University, PO Box 90420, 2024 W. Main St., Durham, NC 27708.

## Abstract

**INTRODUCTION:** Diverse pathogens (viral, bacterial, fungal) have been linked to Alzheimer’s disease (AD) indicating a possibility that the culprit may be compromised immunity rather than particular microbe. If true, then vaccines with broad beneficial effects on immunity might be protective against AD.

**METHODS:** We estimated associations of common adult infections, including herpes simplex, zoster (shingles), pneumonia, and recurrent mycoses, as well as vaccinations against shingles and pneumonia, with the risk of AD in a pseudorandomized sample of the Health and Retirement Study.

**RESULTS:** Shingles, pneumonia, and mycoses diagnosed between ages 65-75, were all associated with higher risk of AD later in life, by 16%-42%. Pneumococcal and shingles vaccines received between ages 65-75 both lowered the risk of AD, by 15%-21%.

**DISCUSSION:** Our results support the idea that the connection between AD and infections involves compromised immunity rather than specific pathogen. We discuss mechanisms by which the declining immune surveillance may promote AD, and the role of biological aging in it. Repurposing of vaccines with broad beneficial effects on immunity could be a reasonable approach to AD prevention. Pneumococcal and zoster vaccines are promising candidates for such repurposing.

## 1. INTRODUCTION

Alzheimer’s Disease (AD) is a complex neurodegenerative disorder that may involve more than one etiological pathway. Understanding the heterogeneity of AD is crucial for development of the efficient AD prevention and treatment. Growing evidence suggests that infections may play an important role in AD development. Very different pathogens (viral, bacterial, fungal) have been linked to AD and related traits in diverse types of studies that until recently more commonly focused on molecular biomarkers and seropositivity than on clinically manifested infections and epidemiological approaches [1-14]. This indicates a possibility that the culprit may not (or not only) be a specific microbe but compromised immunity, which may increase the brain’s vulnerability to a broad range of pathogens in turn contributing to the brain damage and neurodegeneration.

This idea is in line with what we know about major AD risk factors, such as aging, genes in APOE region, brain trauma, and air pollution, which all are associated with weakened immunity and increased vulnerability to infections. For example, AD is strongly correlated with aging, and the aging is accompanied by immunosenescence, manifested in depletion of the body’s immune reserves and slower responses to antigens, leading to the decline in immune surveillance with age. That is, the aging may impair immunity on a broad scale affecting person’s resistance and resilience to various infections [15]. Genetic risk factors for AD, such as SNPs in APOE and TOMM40-NECTIN2 genes, may also be involved in the vulnerability to infections [16]. And so may brain trauma and air pollution (external AD risk factors). E.g., chronic exposure to the traffic-generated air pollution can compromise the blood–brain barrier (BBB) and enhance its permeability to damaging factors, including the infection-related. A traumatic brain injury (TBI) may also affect BBB integrity and immune function, resulting in slower repairs of blood vessels and higher damage to the brain, especially when accompanied by a viral infection [17-19].

If the compromised immunity is a major player in the connection between infections and AD, then interventions aiming to improve the immunity on a broad scale might help delay AD onset. Vaccination is an effective approach to strengthening the immunity and making individuals more resistant and resilient to infections. Recent data has demonstrated that some vaccines (e.g., against tuberculosis (BCG), shingles, flu, and pneumonia) may have broader than expected heterologous (non-specific) off-target effects on health and survival traits that span beyond the protection against target disease [20-34]. Such vaccines may hold a potential for repurposing for AD prevention. For instance, BCG has been sown to lower the AD risk in bladder cancer patients [27, 32, 33]. Identifying the beneficial off-target effects of other vaccines on AD may yield new candidates for repurposing for AD prevention and help clarify the role of the weakened immunity in AD and other health disorders.

In this paper, we address the following questions: Do different kinds of infectious diseases (viral, bacterial, fungal) similarly increase risks of AD and other dementias? Do different adult vaccinations, such as against shingles and pneumonia, similarly reduce risks of AD and other dementias later in life? Answering these questions is essential for clarifying the relationships between infections and AD, and understanding the potential of existing vaccines for repurposing for AD prevention.

## 2. MATERIALS AND METHODS

### 2.1 Data

We used existing data from the Health and Retirement Study (HRS) linked to Medicare administrative claims covering the 1991-2012 period. The HRS is an ongoing national longitudinal dataset that has been fielded every other year since 1992. It collects data on a battery of demographic and socioeconomic characteristics as well as health status and several relevant behavioral factors. The HRS Medicare component provides a continuous individual-level record of all episodes of care paid for by either Medicare Part A (inpatient service) or Medicare Part B (professional services) including data on diagnoses (International Classification of Disease 9^th^ Edition (ICD-9)) and medical procedures (Current Procedural Terminology 4^th^ Edition (CPT-4)), as well as a supplementary source of mortality and demographic information.

### 2.2 ICD-9-CM diagnostic codes used in this study

Using previously validated algorithms [35] and the ICD-9-based definitions presented in Table 1, we identified the probable dates of onset of AD, herpes simplex (HSV), herpes zoster (HZV, shingles), pneumonia, and recurrent mycoses, as well as all vaccinations against herpes zoster (shingles) and pneumonia, with or without an accompanying flu shot. The main ICD-9 diagnostic code for AD was 331.0. However, to account for the fact that AD may co-exists with, or be mistakenly diagnosed as a different type of dementia, and that throughout the study period the ability of physicians to make a verified AD diagnosis varied, we also formed an ‘expanded’ group of the dementia-related diagnoses that included codes for *AD-related dementias* (ADRD), such as frontotemporal, Lewy body, and vascular (331.1, 331.82, 290.4x), as well as some other dementia-related codes - for presenile and senile dementias and psychotic conditions, unspecified cerebral degeneration, senile degeneration of the brain, dementias in conditions classified elsewhere, etc. This ‘expanded’ group of the dementia-related diagnoses is shown as “*ADRD and other dementias (ADRD+)*” in Table1. In our analysis, we used AD diagnostic code (331.0) separately, as well as in combination with the codes in ADRD+ group.

**Table 1.** ICD-9-CM Codes for Primary Outcomes and Predictors.

| Condition | ICD9-CM-Codes |
| --- | --- |
| *Alzheimer's Disease (AD)* | 331.0x |
| *AD-related dementias (ADRD): frontotemporal, Lewy body and vascular* | 331.1x, 331.82, 290.4x |
| *ADRD and other dementias (ADRD+)* | 331.1x, 331.82, 331.2x, 331.9x, 290.xx, 294.xx |
| <sup>1</sup>*Herpes zoster (HZV, shingles)* | 053.xx |
| *Herpes simplex (HSV)* | 054.xx |
| *Herpes all (HSV, HZV, <sup>2</sup>varicella-zoster)* | 052.xx, 053.xx, 054.xx |
| *Pneumonia* | 480.xx-486.xx, 487.0x |
| <sup>3</sup>*Fungal infections (mycoses)* | 110.xx-118.xx |
| <sup>4</sup>*Vaccination against shingles+* | V04.89, V05.8x, V05.4x |
| <sup>5</sup>*Vaccination against pneumonia+* | V06.6x, V03.82 |

It is important to emphasize that in this study we intentionally focused on diagnosed (i.e., *clinically manifested*) infections rather than on seropositivity. The rationale for such focus is the following. Plethora of evidence indicates that prevalence of viral markers in serum and other body fluids is high, so that more than 80% people carry at least one common virus (e.g., herpes simplex or zoster) [36]. This suggests that seropositivity, on its own, would unlikely be the strongest predictor of AD onset. In contrast, clinically manifested infections (new, as well as re-activated) indicate compromised immunity. Indeed, most infectious agents quietly reside in their hosts for decades and become reactivated and symptomatic only in a fraction of population, as result of weakened immune surveillance. The latter may happen due to, e.g., i) damage caused by brain trauma, air pollution, or other harmful exposures that may tap into immune reserves; ii) genetic variation associated with the lower immune resilience; iii) immunosenescence, which is characterized by thymic involution, depletion of the immune reserve, and slower responses to antigens, resulting in decline in the immune surveillance with age [15]. We, therefore, focused on symptomatic disease because it could be a better indicator of weakened immunity and increased host *vulnerability* to infections than seropositivity.

### 2.3 Study sample

We implemented the following sample selection restrictions. Since age is the most important non-genetic risk factor for AD, we set the baseline age for this study at 75. Therefore, individuals who were diagnosed with AD or ADRD+ prior to that age, or individuals who entered the data at more advanced ages were dropped. Then the first and last month for which a Medicare beneficiary had coverage under traditional Medicare (TM) were identified and month-by-month Medicare enrollment data was used to calculate the proportion of time an individual was covered by TM over that period. Individuals covered less than 75% of their total stay in the data were dropped. This was necessary as alternatives to TM do not share their claims data and inclusion of such individuals would introduce bias as there would be no way to identify the presence of a diagnosis. After these restrictions, the final sample sizes were 6,001 for the AD sample, 5,548 for the AD/ADRD+ sample, and 6,171 for the death sample.

### 2.4 Pseudorandomization algorithm

A total of 32 survey-based demographic and socioeconomic characteristics and 30 possible Elixhauser Index-based [37] (other neurological disorders excluded) co-morbidities identified in Medicare records (Supplementary Table A.1) were used in stepwise logistic regressions to predict the probability of an individual having a history of each infectious disease/vaccination by age 75. Individual-level inverse-probability weights (IPW) were created as the reciprocal of the probability to have actually observed a history of an infectious disease/vaccination by age 75. The use of these weights ensures that the *case (*e.g. with infection/vaccine) and *control* (e.g. without infection/vaccine) groups differ only by the presence of the predictor (e.g. medical history of infection/vaccine by age 75), not by other observed factors[38]. Stepwise variable selection was used only in the treatment model for the calculation of propensity scores and evaluation of individual IPWs. The stepwise procedure excluded insignificant predictors which, in turn, have negligible likelihood estimates (i.e., the values of the betas) and therefore have minimal effect on the resulting propensity score and individual weight.

All survey-based measures were defined at the interview nearest to age 75. We included dichotomous variables for three age cohorts based on the design of the HRS – born before 1924, between 1924 and 1930 and between 1942 and 1959 (born between 1930 and 1942 is the comparison group). In addition, we included male gender, black and other (including Hispanics of both skin colors) race, married, three levels of education (less than high school, some college/undergraduate degree, graduate college degree) and veteran status. Variables representing socioeconomic status were being in the lowest (≤$15,579.95) and highest ($43,072.03≤) income quartile (all dollar amounts are inflation adjusted 2010 dollars), the lowest (≤$54,638.95) and highest ($418,463.03≤) wealth quartile, having more than one or more additional health insurance plans, having a life insurance plan, having a long-term care insurance plan, and receiving Social Security Disability Insurance benefits. Factors representing health-related behavior were being underweight (BMI<18.5), overweight (25.0≤BMI≤29.9) or obese (30.0≤BMI), being an occasional (<7 drinks peer week) or heavy (>=7 drinks per week) drinker, smoking at time of interview, and having quit smoking. The final set of survey variables were designed to represent health status not specific to the presence of a disease. These were excellent/very good self-reported health, fair/poor self-reported health, one or two limitations in activities of daily living (ADL: bath, dress, eat, get out of bed, walk across room), three or more ADL limitations, and one or more limitations in instrumental activities of daily living (IADL: phone, money, medications).

### 2.5 Fine-Gray competing risk model

We used the Fine-Gray (1999) [39, 40] competing risk model (with death as the competing risk) for the analysis of AD and AD/ADRD+. This model allows for estimating the relative risk of AD in the AD-free group of living subjects who can die without getting AD or AD/ADRD+. This model was chosen due to the epidemiological characteristics of the diseases under study and the age of the sample. AD is characterized by an advanced age of onset, so the likelihood of dying prior to having a chance to be diagnosed is high. In this case the traditional Cox proportional hazards model, which treats individuals that died as independently censored (and therefore still at risk) is not appropriate. The Fine-Gray model treats the alternative outcome (death, in this study) as a competing risk, and removes deceased individuals from the group of AD-free subjects after death happened. This makes the estimates of AD and AD/ADRD+ risks both more conservative and more realistic. We use the Cox model for the analysis of death, where competing risk is not an issue. Finally, since the use of individual level weights acts to make independent observations related to each other (e.g., a single individual, before IPW represents a number of individuals equal to his/her weight after IPW) we used the robust sandwich variance estimate to calculate the confidence intervals of our weighted models.

### 2.6 Proportions of targeted events in study population

The study-wide proportion of adverse events (a.k.a. failures) was 11.8% for AD, 27.4% for AD/ADRD+, and 36.5% for death. Summary statistics for all three study subsamples with respect to the primary outcomes and predictors of interest as well as all variables used in the pseudorandomization algorithm are presented in Supplementary Table A.2. The sample subgroups were followed for an average of 6.08 (AD), 5.69 (AD/ADRD+) and 6.33 (Death) years until exit. The lowest average follow-up time for AD/ADRD+ is consistent with the wider selection of diagnoses codes used for this group. The average time since the latest HRS interview was about 0.77 years, which is excellent given that HRS is a bi-annual survey. About 11% of the population had a history of the herpes virus at baseline. The majority (9%) of this was accounted by herpes/zoster with herpes/simplex present in only 3% of the samples. About 19-21% of the sample had a history of pneumonia and 14-16% of a fungal infection. Levels of herpes vaccinations between ages 65 and 75 were low at about 4%; however, pneumonia vaccinations were much higher at about 29-30%. Supplementary Table A.3 demonstrates the overlap between our primary outcomes and main explanatory variables before and after pseudorandomization. With the possible exception of herpes simplex and shingles vaccine there is sufficient overlap even before pseudorandomization. After pseudorandomization all categories are fairly evenly matched. The pseudorandomization quality for all models was excellent (Supplementary Tables A.4 - A.6). This can be seen by the lack of statistically significant differences between the case and control groups after the pseudorandomization procedure.

## 3. RESULTS

Table 2 shows results of our analysis using two models: (i) the *univariate* model that uses only the primary predictor without pseudorandomization-based weights or co-variates in a Fine-Gray competing risk model (Cox proportional hazards model for mortality); (ii) the *pseudorandomized* model that uses the inverse-probability weights in a Fine-Gray competing risk model (Cox proportional hazards model for mortality). One can see from the Table 2 that a herpes virus infection (herpes simplex or zoster) diagnosed between ages 65 and 75 is associated with an about 25% increase in the risk of AD onset after age 75, and the effect becomes significant after pseudorandomization (Hazard Ratio [HR]:1.25; 95% Confidence Interval [CI]:1.08-1.44). Separately, only herpes zoster (shingles) showed a statistically significant association with AD (HR:1.23; CI:1.06-1.41). Both, simplex and zoster infections were not associated with AD/ADRD+, or with all-cause mortality risk once population heterogeneity was accounted for through pseudorandomization. Pneumonia diagnosis was statistically significantly associated with increased risks of AD (HR:1.16; CI:1.001-1.33), AD/ADRD+ (HR:1.12; CI:1.02-1.24) and death (HR:1.16; CI:1.03-1.30]. The largest effect sizes were observed for the associations between recurrent fungal infections (mycoses) and AD (HR:1.42; CI:1.24-1.63), and AD/ADRD+ (HR:1.44; CI:1.31-1.58), in pseudorandomization model. Notably, the association between mycoses and death that was seen in univariate model (HR:1.31; CI:1.17-1.46) disappeared after pseudorandomization (HR:1.00; CI:0.88-1.14).

**Table 2.** Effects of Vaccines and Infections on Risk of Alzheimer’s Disease, Other Dementias, and All-cause Mortality.

| Predictor<br>(Infection or Vaccine) | Competing-Risk Univariate, Without PSM |  |  | Competing-Risk Pseudorandomized<br>PSM, Fine-Gray |  |  |
| --- | --- | --- | --- | --- | --- | --- |
|  | AD | AD/ADRD+ | Death<sup>1</sup> | AD | AD/ADRD+ | Death<sup>1</sup> |
| *Herpes (HSV, HZV/*<br>*varicella-zoster)* | 1.24<br>[ 0.98- 1.56] | 1.08<br>[ 0.91- 1.27] | 1.14<br>[ 1.00- 1.31] | **1.25\*\***<br>[ 1.08- 1.44] | 1.10<br>[ 1.00- 1.21] | 1.09<br>[ 0.94- 1.27] |
| *Herpes zoster (HZV,*<br>*shingles)* | 1.27<br>[ 0.99- 1.63] | 1.08<br>[ 0.90- 1.30] | 1.16*<br>[ 1.00- 1.34] | **1.23\*\***<br>[ 1.06- 1.41] | 1.09<br>[ 0.99- 1.20] | 1.09<br>[ 0.93- 1.28] |
| *Herpes simplex (HSV)* | 1.17<br>[ 0.73- 1.89] | 1.13<br>[ 0.81- 1.59] | 1.14<br>[ 0.87- 1.51] | 0.99<br>[ 0.86- 1.15] | 1.08<br>[ 0.98- 1.19] | 1.17<br>[ 0.85- 1.61] |
| *Pneumonia* | 1.08<br>[ 0.90- 1.29] | 1.16*<br>[ 1.02- 1.31] | 1.82**<br>[ 1.66- 2.00] | **1.16\***<br>[ 1.00- 1.33] | **1.12\***<br>[ 1.02- 1.24] | **1.16\***<br>[ 1.03- 1.30] |
| *Recurrent fungal infection*<br>*(mycoses)* | 1.35**<br>[ 1.11- 1.63] | 1.42**<br>[ 1.24- 1.62] | 1.31**<br>[ 1.17- 1.46] | **1.42\*\***<br>[ 1.24- 1.63] | **1.44\*\***<br>[ 1.31- 1.58] | 1.00<br>[ 0.88- 1.14] |
| *Vaccination against*<br>*shingles (herpes zoster)* | 0.91<br>[ 0.60- 1.40] | 0.94<br>[ 0.71- 1.25] | 0.72*<br>[ 0.55- 0.94] | **0.79\*\***<br>[ 0.68- 0.93] | **0.90\***<br>[ 0.81- 0.99] | 0.80<br>[ 0.55- 1.17] |
| *Vaccination against*<br>*pneumonia* | 0.79*<br>[ 0.66- 0.96] | 0.87*<br>[ 0.77- 0.99] | 0.93<br>[ 0.84- 1.03] | **0.85\***<br>[ 0.73- 0.99] | **0.88\***<br>[ 0.79- 0.97] | **0.85\***<br>[ 0.74- 0.97] |
| *N* | 6,001 | 5,548 | 6,171 | 6,001 | 5,548 | 6,171 |
| *N Primary Outcome* | 706 | 1,520 | 2,250 | **706** | **1,520** | **2,250** |
| *N Competing Risk (Death)* | 1,738 | 1,111 |  | 1,738 | 1,111 |  |

<sup>1</sup> The Cox proportional hazards model was used for death in the absence of a competing risk.
\*p<0.05; \*\*p<0.01

Vaccination against shingles with the live attenuated virus (Zostavax) was strongly associated with lower risk of AD (HR:0.79; CI:0.68-0.93), and less prominently (but still statistically significantly) with the lower risk of AD/ADRD+ (HR:0.90; CI:0.81-0.99) after pseudorandomization but not before. A protective effect of this vaccine on all-cause mortality risk seen before pseudorandomization became not significant after accounting for population heterogeneity. Vaccination against pneumonia was associated with statistically significantly lower risks of AD (HR:0.85; CI:0.73-0.99), AD/ADRD+ (HR:0.88; CI:0.79-0.97), and death (HR:0.85; CI:0.74-0.97) (Table 2).

## 4. DISCUSSION

In our study, shingles, pneumonia, and fungal infections were all associated with significantly higher risk of AD onset in a pseudorandomized sample of HRS participants, while pneumonia and shingles (zoster) vaccines were both protective against AD (Table 2). These results are in line with the idea that AD is linked to compromised immunity rather than to a specific pathogen. The effects of infections and vaccines were overall stronger for AD than for other dementias in this study, suggesting that immune system may play more important role in AD than in the other dementias, which deserves further investigation.

Our results indicate that shingles and pneumonia vaccines are promising candidates for repurposing for AD prevention. Mechanisms by which the existing vaccines could provide off-target protection against AD may involve both non-specific and specific effects of these vaccines, including: (i) training of the innate immunity that may prolong active life of macrophages; (ii) production of lymphocytes with cross-reactivity that may enhance adaptive immune responses, and (iii) reduction in the risk of target disease per se that otherwise might contribute to neurodegeneration, among other factors [22, 41, 42].

Below, we discuss these and other potential mechanisms of the observed associations in more detail.

### 4.1. Aging may contribute to the association between infections and AD by weakening the immune surveillance

Age is the strongest risk factor for AD, and it is also associated with weakening the immunity. Indeed, biological aging is characterized by immunosenescence, slower responses to antigens, and compromised BBB integrity, which results in declining immune surveillance with age [15, 43], allowing for re-activation of latent microbes (e.g., varicella zoster virus) and increasing their chances of reaching the brain and causing inflammation and neuronal death. Aging may also be a reason why different microorganisms seem to play similar roles in AD. This could be because the aging-related changes fundamentally affect the body and may exacerbate detrimental effects of various pathogens and other factors. For instance, the aging-related impairment in myelin maintenance and debris clearance by microglia may facilitate axon myelin depletion caused by different stressors [44-47].

### 4.2 Could beneficial effects of vaccines on AD be due to preventing the original target disease?

Vaccines may potentially contribute to a reduction in AD risk by preventing cases of the originally targeted disease. E.g., encephalitis was described as a complication in herpes infections, and was associated with cognitive impairment [14] [48] [49], so preventing the herpes infections with vaccines could reduce cases of the encephalitis that otherwise would contribute to AD or other dementias. However, studies that reported a lower incidence of dementia after vaccination against shingles suggested that reduction of shingles cases is not likely the mechanism by which zoster vaccine lowers dementia risk [50-52], because the beneficial effect of vaccine on dementia was seen in people with, as well as without, history of shingles [50, 51]. Other studies that reported the off-target effects of vaccines on health traits also concluded that such effects cannot be explained by the pathogen-specific immune protection that these vaccines offer, suggesting that *non-specific* immune system modulation by the vaccines may be a widespread phenomenon [53, 54].

### 4.3 Strengthening the immunity broadly may explain the off-target effects of vaccines

If some vaccines induce *non-specific* immune responses that can boost immunity broadly, then what are mechanisms of such effects? [22, 26]. While pathogen-specific mechanisms of vaccines have been studied for decades, the biological machinery behind their non-specific effects (beneficial or detrimental to health) remains underexplored [26]. The tuberculosis Bacillus Calmette-Guérin (BCG) vaccine has been most intensively studied in this regard [23, 25, 31, 55]. Several studies suggested that vaccines with beneficial off-target effects can produce non-specific immune responses that improve the overall immune robustness and resilience as a result of *trained immunity* involving the process of priming macrophages that leads to their prolonged functioning. The protective off-target effect may also be due to the *antigen cross-reactivity* and *heterologous lymphocyte activation* [22, 28, 56]. For example, both zoster and pneumococcal vaccines were shown to broadly stimulate CD4 T cell responses. The CD4 receptor is found on the surface of many immune cells, including T helpers, monocytes, macrophages, and dendritic cells, so it can be involved in a wide range of infections. The decline in CD4 T cells responsiveness has been implicated in AD and neuronal damage in infectious diseases. Therefore, one mechanism by which such vaccines might provide protection against AD could involve broad boosting the CD4 T cell responses [57-59].

In our study, shingles and pneumonia vaccines had beneficial effects on AD, as well as (though less prominently) on all-cause mortality, which may reflect the non-specific effects of these vaccines on immunity. Other studies, using different data, also reported a lower incidence of dementia following the shingles vaccination, which is in agreement with our results [50, 51]. Schnier et al. (2022) concluded that the protective effect of shingles vaccine on dementia risk in their study was not a consequence of shingles reduction, but was likely due to non-specific effects of this vaccine on immunity. It was also suggested that COVID-19 may suppress immunity and trigger re-activation of latent herpes zoster [60]. This indicates that shingles vaccine might provide protection against AD in COVID-19 patients, which deserves further investigation. Several other studies also supported beneficial off-target effects of pneumonia vaccine (alone, or in combination with a flu shot) on health/survival traits, including on preventing COVID-19 [21, 24, 61, 62].

### 4.4 Limitations

#### Live and non-live vaccines may differ in their effects on AD

The HRS data used in this study cover the 1991-2012 period, during which the only shingles vaccine available was the *live* attenuated zoster vaccine (Zostavax) approved in 2006. Starting 2017, a new recombinant adjuvanted zoster vaccine (Shingrix) has been in use, while Zostavax has not been used since October 2020. This change in vaccine type (from *live* to *non-live*) may affect replication of the observed association between shingles vaccine and lower AD risk, because it is unclear if the *non-live* vaccine has the same beneficial effect on AD, as the *live* vaccine. Several studies explored similar situation for some other vaccines though. In a study of childhood vaccinations, only *live* attenuated vaccines showed beneficial heterologous effects on health/survival traits, while *non-live* vaccines did not [23, 25, 63]. Though similar differences have yet to be demonstrated for *live* vs. *non-live* shingles vaccine, the possibility exists that only *live* attenuated zoster vaccine (used in our analysis) reduces AD risk, while the recombinant vaccine does not. Comparing the live and *non-live* zoster vaccines for their effects on AD is, therefore, important next step of evaluating these vaccines for repurposing for AD prevention.

The following remarks about interpretation of this study results should also be made. First, the beneficial effect of the *live* shingles vaccine on the risk of AD should not be generalized to the *non-live* recombinant vaccine, which is currently in use. Second, vaccination against shingles is covered by Medicare Part D (prescription drug coverage). Therefore, individuals who have only Medicare parts A & B would pay for this vaccination either out-of-pocket or through a secondary health insurance. This leads to our comparison group (without shingles vaccine) being a mix of individuals who a) did not receive a vaccine, and b) who obtained the vaccine through a non-Medicare payer. This means that our result (∼21% reduction in AD risk following the vaccination against shingles) represents a *lower* estimate of the AD-preventive effect of this vaccine.

#### Medicare claims data share other well-known limitations of administrative data

Although the majority of older U.S. adults aged 65+ are eligible for Medicare, the system does not have 100% coverage, and may miss out some specific at-risk groups. Also, data are only available for services provided by medical offices, so individuals who choose not to go to the doctor, or pay out of the pocket, cannot be accounted for in this analysis.

***A (potential) limitation is that herpesviruses are common inhabitants of human body*** and can be found in latent forms in the majority of the population [64], so it may seem unlikely that such viruses would show any substantial effect on AD. Indeed, herpesviruses (as well as many other microbes) may quietly reside in their hosts for decades, or even for a lifetime, without causing harm, indicating that in such cases the immune system can successfully control these pathogens. These viruses become active and may cause clinical symptoms only in certain individuals, an indication that their immune surveillance is impaired and allows those viruses to proliferate. In other words, the reactivation and clinical manifestation of a latent viral infection is a sign of weakened immunity, which can play a role in AD [6]. This is why we used diagnosed (i.e., *clinically manifested*) cases of the infection in this study, rather than seropositivity, as an indicator of the compromised immunity and increased host vulnerability to various infections.

## 5. CONCLUSION

Various pathogens (viral, bacterial, fungal) have been associated with Alzheimer’s disease in recent and past research, suggesting a possibility that weakened host immunity (rather than specific microbe) plays a major role in etiology of this disease through the increased brain’s vulnerability to many microorganisms and toxins, in turn contributing to neurodegeneration.

Our study found that clinically manifested herpes zoster (shingles), pneumonia, and recurrent fungal infections are *all* associated with increased AD risk, which supports the idea that the mechanism connecting AD and infections involves compromised immunity rather than particular pathogen.

We also found that shingles and pneumonia vaccines have beneficial off-target effects on AD and might be protective against it’s development. Potential mechanisms may involve boosting the non-specific immune response and resilience as the result of the antigen cross-reactivity, heterologous lymphocyte activation, and trained innate immunity [22, 28, 56], among other factors. We conclude that the live attenuated zoster vaccine and pneumococcal polysaccharide vaccine are promising candidates for repurposing for AD prevention in older adults. It remains to be determined whether a non-live shingles vaccine has a similar beneficial effect on AD.

## Data Availability

Data subject to third party restrictions. The data that support the findings of this study are available from the Health and Retirement Study (HRS), sponsored by the National Institute on Aging (grant number U01-AG009740) and led by the University of Michigan. Restrictions apply to the availability of these data, which were used under license for this study. Data are available at https://hrs.isr.umich.edu with the permission of the HRS.

## Supplementary Materials

The Supplemental Tables S1-S6.

## Author Contributions

Conceptualization, S.U. and A.P.Y.; writing, original draft preparation, S.U. and A.P.Y.; review, editing, updates, I.A., E.S., K.A., A.Y.; data preparation, statistical analysis, A.P.Y., G.G., H.D. All authors have read and agreed to the final version of the manuscript.

## Funding

Research reported in this publication was supported by the National Institutes of Aging of the National Institutes of Health (NIA/NIH) under award numbers RF1AG046860, R01AG076019, R01AG062623, R01AG066133, and R01AG063971.

## Acknowledgements

We acknowledge investigators and participants of the Health and Retirement Study, sponsored by the National Institute on Aging (grant number U01-AG009740) and led by the University of Michigan.

## Conflicts of Interests

The authors declare that they have no conflicts of interests.

## CONSENT STATEMENT

A waiver of all elements of informed consent listed in 45CFR46.116(a)(1–8) and 45CFR46.116 (b)(1–6); and the HIPAA core elements in 45CFR164.508(c)(1)(i–vi) and the required statements in 45CFR164.508(c)(2)(i–iii) was obtained from the DUHS Institutional Review Board (Duke University), Federalwide Assurance No: FWA 00009025, to conduct secondary analyses of the data under fund codes RF1AG046860, R01AG076019, R01AG062623, R01AG066133, and R01AG063971. The rationale for each is that the waiver is essential for this research to be feasible. The control over the informed consent procedures is regulated by the University of Michigan Institutional Review Board, Federal wide Assurance No: FWA 00004969, under the UM Health Sciences/Behavioral Sciences IRB Protocol: HUM00061128 approved through 10/18/2018 and associated protocols: HUM00056464, HUM00002562, HUM00074501, HUM00079949, HUM00080925, HUM00085942, HUM00099822, HUM00103072, HUM00106904, HUM00122335, REP00000046. Prior to each interview, participants are provided with a written informed consent information document. At the start of each interview, all respondents are read a confidentiality statement, and give oral consent by agreeing to do the interview.

## Notes

### Competing Interest Statement

The authors have declared no competing interest.

### Author Declarations

The Duke University Health System Institutional Review Board for Clinical Investigations, Federalwide Assurance No: FWA 00009025 gave ethical approval for this work IRB ID: Pro00048443, Pro00109279, Pro00105166, Pro00103556, Pro00006711.

