## Supplemental Tables 1-6 for "Associations of infections and vaccines with Alzheimer’s disease point to a major role of compromised immunity rather than specific pathogen in AD"

| **Supplementary Table 1. ICD-9-CM Codes Used in the Pseudorandomization Model** | |
| --- | --- |
| **Condition** | **ICD9-CM-Codes** |
| **Outcomes** | |
| *Alzheimer's Disease (AD)* | 331.0x |
| *Alzheimer's Disease and other dementias (AD/ADRD+)* | 331.0x 331.1x 331.2x 331.9x 331.82 290.xx 294.xx |
| **Primary Explanatory Variables** | |
| *Herpes all (HSV, HZV, varicella-zoster)* | 052.xx 053.xx 054.xx |
| *Herpes zoster (HZV, shingles)* | 053.xx |
| *Herpes simplex (HSV)* | 054.xx |
| *Pneumonia* | 480.xx-486.xx 487.0x |
| *Recurrent fungal infection (mycoses)* | 110.xx-118.xx |
| *Vaccination against shingles* | V05.4x V04.89 V05.8x |
| *Vaccination against pneumonia* | V06.6x V03.82 |
| **Elixhauser Index-based Co-morbidities Used in Pseudorandomization Algorithm** | |
| *Congestive heart failure* | 398.91 402.01 402.11.402.91 404.01 404.03 404.11 404.13 404.91 404.93 425.4-425.9 428.x |
| *Cardiac arrhythmias* | 426.0 426.13 426.7 426.9 426.10 426.12 427.0-427.4 427.6-427.9 785.0 996.01 996.04 V45.0 V53.3 |
| *Valvular disease* | 093.2 394.x-397.x 424.x 746.3-746.6 V42.2 V43.3 |
| *Pulmonary circulation Disorders* | 415.0 415.1 416.x 417.0 417.8 417.9 |
| *Peripheral vascular disorders* | 093.0 437.3 440.x 441.x 443.1-443.9 447.1 557.1 557.9 V43.4 |
| *Hypertension, uncomplicated* | 401.x |
| *Hypertension, complicated* | 402.x-405.x |
| *Paralysis* | 334.1 342.x 343.x 344.0-344.6 344.9 |
| *Chronic pulmonary disease* | 416.8 416.9 490.x-505.x 506.4 508.1 508.8 |
| *Diabetes, uncomplicated* | 250.0-250.3 |
| *Diabetes, complicated* | 250.4-250.9 |
| *Hypothyroidism* | 240.9 243.x 244.x 246.1 246.8 |
| *Renal failure* | 403.01 403.11 403.91 404.02 404.03 404.12 404.13 404.92 404.93 585.x 586.x 588.0 V42.0 V45.1 V56.x |
| *Liver disease* | 070.22 070.23 070.32 070.33 070.44 070.54 070.6 070.9 456.0-456.2 570.x 571.x 572.2-572.8 573.3 573.4 573.8 573.9 V42.7 |
| *Peptic ulcer disease excluding bleeding* | 531.7 531.9 532.7 532.9 533.7 533.9 534.7 534.9 |
| *AIDS/H1V* | 042.x-044.x |
| *Lymphoma* | 200.x-202.x 203.0 238.6 |
| *Metastatic cancer* | 196.x-199.x |
| *Solid tumor without metastasis* | 140.x-172.x 174.x-195.x |
| *Rheumatoid arthritis/ collagen vascular diseases* | 446.x 701.0 710.0-710.4 710.8 710.9 711.2 714.x 719.3 720.x 725.x 728.5 728.89 729.30 |
| *Coagulopathy* | 286.x 287.1 287.3-287.5 |
| *Obesity* | 278.0 |
| *Weight loss* | 260.x-263.x 783.2 799.4 |
| *Fluid and electrolyte disorders* | 253.6 276.x |
| *Blood loss anemia* | 280.0 |
| *Deficiency anemia* | 280.1-280.9 281.x |
| *Alcohol abuse* | 265.2 291.1-291.3 291.5-291.9 303.0 303.9 305.0 357.5 425.5 535.3 571.0-571.3 980.x V11.3 |
| *Drug abuse* | 292.x 304.x 305.2-305.9 V65.42 |
| *Psychoses* | 293.8 295.x 296.04 296.14 296.44 296.54 297.x 298.x |
| *Depression* | 296.2 296.3 296.5 300.4 309.x 311 |

| **Supplementary Table 2. Summary Statistics** | | | |
| --- | --- | --- | --- |
|  | **AD** | **AD/ADRD+** | **Death** |
| *Follow-up in years* | 6.08 (4.23) | 5.69 (4.06) | 6.33 (4.34) |
| *`* | 0.77 (1.18) | 0.77 (1.18) | 0.78 (1.18) |
| *Herpes all (HSV, HZV, varicella-zoster)* | 0.11 (0.31) | 0.11 (0.31) | 0.11 (0.31) |
| *Herpes zoster (HZV, shingles)* | 0.09 (0.28) | 0.09 (0.28) | 0.09 (0.28) |
| *Herpes simplex (HSV)* | 0.03 (0.16) | 0.03 (0.16) | 0.03 (0.16) |
| *Pneumonia* | 0.20 (0.40) | 0.19 (0.39) | 0.21 (0.40) |
| *Recurrent fungal infection (mycoses)* | 0.15 (0.36) | 0.14 (0.35) | 0.16 (0.36) |
| *Vaccination against shingles* | 0.04 (0.19) | 0.04 (0.19) | 0.04 (0.19) |
| *Vaccination against pneumonia* | 0.30 (0.46) | 0.29 (0.46) | 0.30 (0.46) |
| *Born before 1924* | 0.26 (0.44) | 0.27 (0.44) | 0.26 (0.44) |
| *Born 1924 - 1930* | 0.25 (0.43) | 0.25 (0.43) | 0.25 (0.43) |
| *Born 1942 - 1959* | 0.01 (0.07) | 0.01 (0.07) | 0.01 (0.07) |
| *Male* | 0.45 (0.50) | 0.45 (0.50) | 0.45 (0.50) |
| *Black* | 0.11 (0.32) | 0.11 (0.31) | 0.12 (0.32) |
| *Hispanic* | 0.05 (0.22) | 0.05 (0.22) | 0.05 (0.23) |
| *Other race* | 0.02 (0.14) | 0.02 (0.14) | 0.02 (0.14) |
| *Married* | 0.64 (0.48) | 0.64 (0.48) | 0.63 (0.48) |
| *Education: less than high-school* | 0.27 (0.44) | 0.26 (0.44) | 0.27 (0.44) |
| *Education: some college* | 0.14 (0.34) | 0.14 (0.35) | 0.13 (0.34) |
| *Education: graduate degree* | 0.07 (0.26) | 0.08 (0.26) | 0.07 (0.26) |
| *Veteran* | 0.33 (0.47) | 0.33 (0.47) | 0.32 (0.47) |
| *Receives SSDI benefits* | 0.02 (0.13) | 0.01 (0.12) | 0.02 (0.13) |
| *Lowest income quartile* | 0.22 (0.42) | 0.22 (0.41) | 0.23 (0.42) |
| *Highest income quartile* | 0.25 (0.43) | 0.25 (0.43) | 0.24 (0.43) |
| *Lowest wealth quartile* | 0.22 (0.41) | 0.20 (0.40) | 0.22 (0.42) |
| *Highest wealth quartile* | 0.27 (0.44) | 0.27 (0.45) | 0.26 (0.44) |
| *One or more health insurance plans* | 0.03 (0.17) | 0.03 (0.17) | 0.03 (0.17) |
| *Life insurance* | 0.65 (0.48) | 0.65 (0.48) | 0.65 (0.48) |
| *Long-term care insurance* | 0.15 (0.36) | 0.15 (0.36) | 0.15 (0.35) |
| *Smokes* | 0.09 (0.29) | 0.09 (0.28) | 0.09 (0.29) |
| *Quit smoking* | 0.49 (0.50) | 0.49 (0.50) | 0.49 (0.50) |
| *Occasional drinker* | 0.21 (0.41) | 0.22 (0.41) | 0.21 (0.41) |
| *Heavy drinker* | 0.08 (0.27) | 0.08 (0.27) | 0.08 (0.27) |
| *Underweight* | 0.02 (0.14) | 0.02 (0.13) | 0.02 (0.14) |
| *Overweight* | 0.41 (0.49) | 0.41 (0.49) | 0.40 (0.49) |
| *Obese* | 0.24 (0.42) | 0.23 (0.42) | 0.23 (0.42) |
| *Excellent/good self-reported health* | 0.31 (0.46) | 0.29 (0.45) | 0.32 (0.47) |
| *Poor/bad self-reported health* | 0.36 (0.48) | 0.38 (0.48) | 0.36 (0.48) |
| *One or two ADL limitations* | 0.12 (0.33) | 0.12 (0.32) | 0.12 (0.33) |
| *Three or more ADL limitations* | 0.05 (0.21) | 0.04 (0.19) | 0.05 (0.23) |
| *One or more IADL limitations* | 0.09 (0.28) | 0.07 (0.25) | 0.10 (0.30) |
| *Congestive heart failure* | 0.14 (0.35) | 0.13 (0.33) | 0.14 (0.35) |
| *Cardiac arrhythmias* | 0.19 (0.39) | 0.18 (0.38) | 0.19 (0.39) |
| *Valvular disease* | 0.05 (0.22) | 0.05 (0.22) | 0.05 (0.22) |
| *Pulmonary circulation Disorders* | 0.03 (0.18) | 0.03 (0.17) | 0.03 (0.18) |
| *Peripheral vascular disorders* | 0.15 (0.36) | 0.14 (0.35) | 0.16 (0.37) |
| *Hypertension, uncomplicated* | 0.63 (0.48) | 0.62 (0.48) | 0.63 (0.48) |
| *Hypertension, complicated* | 0.11 (0.32) | 0.10 (0.30) | 0.12 (0.32) |
| *Paralysis* | 0.04 (0.21) | 0.04 (0.19) | 0.05 (0.21) |
| *Chronic pulmonary disease* | 0.23 (0.42) | 0.22 (0.41) | 0.23 (0.42) |
| *Diabetes, uncomplicated* | 0.24 (0.43) | 0.22 (0.42) | 0.24 (0.43) |
| *Diabetes, complicated* | 0.08 (0.28) | 0.07 (0.26) | 0.09 (0.28) |
| *Hypothyroidism* | 0.15 (0.36) | 0.14 (0.35) | 0.15 (0.36) |
| *Renal failure* | 0.05 (0.22) | 0.04 (0.20) | 0.05 (0.22) |
| *Liver disease* | 0.03 (0.17) | 0.03 (0.17) | 0.03 (0.18) |
| *Peptic ulcer disease excluding bleeding* | 0.04 (0.19) | 0.03 (0.18) | 0.04 (0.19) |
| *AIDS/H1V* | 0.00 (0.03) | 0.00 (0.03) | 0.00 (0.03) |
| *Lymphoma* | 0.01 (0.10) | 0.01 (0.10) | 0.01 (0.10) |
| *Metastatic cancer* | 0.02 (0.14) | 0.02 (0.14) | 0.02 (0.14) |
| *Solid tumor without metastasis* | 0.15 (0.36) | 0.15 (0.36) | 0.15 (0.36) |
| *Rheumatoid arthritis/ collagen vascular diseases* | 0.07 (0.25) | 0.06 (0.25) | 0.07 (0.25) |
| *Coagulopathy* | 0.05 (0.22) | 0.05 (0.21) | 0.05 (0.22) |
| *Obesity* | 0.05 (0.22) | 0.05 (0.21) | 0.05 (0.22) |
| *Weight loss* | 0.05 (0.22) | 0.04 (0.20) | 0.05 (0.22) |
| *Fluid and electrolyte disorders* | 0.13 (0.33) | 0.11 (0.32) | 0.14 (0.34) |
| *Blood loss anemia* | 0.02 (0.13) | 0.01 (0.12) | 0.02 (0.14) |
| *Deficiency anemia* | 0.08 (0.28) | 0.07 (0.26) | 0.08 (0.28) |
| *Alcohol abuse* | 0.01 (0.10) | 0.01 (0.09) | 0.01 (0.11) |
| *Drug abuse* | 0.00 (0.06) | 0.00 (0.05) | 0.00 (0.07) |
| *Psychoses* | 0.01 (0.12) | 0.01 (0.09) | 0.02 (0.14) |
| *Depression* | 0.10 (0.30) | 0.09 (0.28) | 0.11 (0.31) |
| *N* | 6,001 | 5,548 | 6,171 |
| *N Primary Outcome* | 706 | 1,520 | 2,250 |
| *N Competing Risk (Death)* | 1,738 | 1,111 |  |
| Note: Numbers presented are subsample means with standard deviations in parentheses. |  |  |  |

| **Supplementary Table 3. Cross Sample Sizes for Primary Outcomes and Main Explanatory Variables** | | | | | | | | | |
| --- | --- | --- | --- | --- | --- | --- | --- | --- | --- |
|  |  | **AD Sample** | | | **AD/ADRD+ Sample** | | | **Dead Sample** | |
| **Variable** | **Value** | **Censored** | **AD** | **Dead** | **Censored** | **AD/ADRD+** | **Dead** | **Censored** | **Dead** |
| **Panel A. Before Weighting** | | | | | | | | | |
| *Herpes all (HSV, HZV, varicella-zoster)* | 1 | 384 | 82 | 174 | 312 | 156 | 116 | 425 | 231 |
|  | 0 | 3,173 | 624 | 1,564 | 2,605 | 1,364 | 995 | 3,496 | 2,019 |
| *Herpes zoster (HZV, shingles)* | 1 | 308 | 69 | 143 | 250 | 128 | 94 | 341 | 194 |
|  | 0 | 3,249 | 637 | 1,595 | 2,667 | 1,392 | 1,017 | 3,580 | 2,056 |
| *Herpes simplex (HSV)* | 1 | 98 | 18 | 40 | 77 | 37 | 27 | 107 | 51 |
|  | 0 | 3,459 | 688 | 1,698 | 2,840 | 1,483 | 1,084 | 3,814 | 2,199 |
| *Pneumonia* | 1 | 596 | 148 | 466 | 454 | 313 | 290 | 669 | 600 |
|  | 0 | 2,961 | 558 | 1,272 | 2,463 | 1,207 | 821 | 3,252 | 1,650 |
| *Recurrent fungal infection (mycoses)* | 1 | 505 | 125 | 269 | 376 | 255 | 149 | 589 | 368 |
|  | 0 | 3,052 | 581 | 1,469 | 2,541 | 1,265 | 962 | 3,332 | 1,882 |
| *Vaccination against shingles* | 1 | 160 | 21 | 45 | 137 | 46 | 25 | 173 | 56 |
|  | 0 | 3,397 | 685 | 1,693 | 2,780 | 1,474 | 1,086 | 3,748 | 2,194 |
| *Vaccination against pneumonia* | 1 | 1,309 | 135 | 364 | 1,080 | 320 | 235 | 1,404 | 452 |
|  | 0 | 2,248 | 571 | 1,374 | 1,837 | 1,200 | 876 | 2,517 | 1,798 |
| **Panel B. After Weighting^1^** | | | | | | | | | |
| *Herpes all (HSV, HZV, varicella-zoster)* | 1 | 1,731 | 410 | 849 | 1,410 | 799 | 563 | 1,947 | 1,132 |
|  | 0 | 1,790 | 349 | 873 | 1,464 | 758 | 553 | 1,966 | 1,126 |
| *Herpes zoster (HZV, shingles)* | 1 | 1,762 | 402 | 834 | 1,423 | 790 | 560 | 1,947 | 1,135 |
|  | 0 | 1,784 | 348 | 871 | 1,463 | 759 | 553 | 1,966 | 1,123 |
| *Herpes simplex (HSV)* | 1 | 1,806 | 322 | 870 | 1,340 | 777 | 623 | 1,983 | 1,103 |
|  | 0 | 1,779 | 353 | 870 | 1,479 | 768 | 561 | 1,959 | 1,126 |
| *Pneumonia* | 1 | 1,699 | 384 | 897 | 1,392 | 802 | 561 | 1,887 | 1,179 |
|  | 0 | 1,814 | 349 | 857 | 1,488 | 760 | 545 | 1,994 | 1,111 |
| *Recurrent fungal infection (mycoses)* | 1 | 1,698 | 461 | 806 | 1,325 | 961 | 451 | 1,932 | 1,107 |
|  | 0 | 1,796 | 344 | 896 | 1,479 | 753 | 579 | 1,978 | 1,155 |
| *Vaccination against shingles* | 1 | 2,071 | 270 | 732 | 1,782 | 661 | 393 | 2,269 | 872 |
|  | 0 | 1,726 | 347 | 855 | 1,415 | 748 | 549 | 1,915 | 1,116 |
| *Vaccination against pneumonia* | 1 | 1,986 | 279 | 692 | 1,640 | 637 | 465 | 2,189 | 857 |
|  | 0 | 1,749 | 379 | 916 | 1,428 | 803 | 575 | 1,936 | 1,189 |
| 1 Weighted sample size rounded to the nearest person for display purposes. | | | | | | | | | |

| **Supplementary Table 4: Assessment of Pseudorandomization Quality - Alzheimer's Disease*** | | | | | | | |
| --- | --- | --- | --- | --- | --- | --- | --- |
|  |  | **Before Pseudorandomization** | | | **After Pseudorandomization** | | |
|  |  | **Group** | | **χ2** | **Group** | | **χ2** |
| **Infection/Vaccine** | **Variable** | **Control** | **Treatment** | **p-val** | **Control** | **Treatment** | **p-val** |
| *Herpes all (HSV, HZV, varicella-zoster)* | *Diabetes, complicated* | 0.08 | 0.12 | <.01 | 0.08 | 0.09 | 0.44 |
| *Herpes all (HSV, HZV, varicella-zoster)* | *Hypothyroidism* | 0.14 | 0.23 | <.01 | 0.15 | 0.15 | 0.86 |
| *Herpes all (HSV, HZV, varicella-zoster)* | *Lymphoma* | 0.01 | 0.02 | <.01 | 0.01 | 0.01 | 0.87 |
| *Herpes all (HSV, HZV, varicella-zoster)* | *Solid tumor without metastasis* | 0.15 | 0.18 | 0.02 | 0.15 | 0.16 | 0.67 |
| *Herpes all (HSV, HZV, varicella-zoster)* | *Rheumatoid arthritis/ collagen vascular diseases* | 0.06 | 0.12 | <.01 | 0.07 | 0.07 | 0.87 |
| *Herpes all (HSV, HZV, varicella-zoster)* | *Depression* | 0.09 | 0.15 | <.01 | 0.10 | 0.11 | 0.66 |
| *Herpes all (HSV, HZV, varicella-zoster)* | *Born before 1924* | 0.27 | 0.21 | <.01 | 0.26 | 0.26 | 0.71 |
| *Herpes all (HSV, HZV, varicella-zoster)* | *Male* | 0.46 | 0.38 | <.01 | 0.45 | 0.45 | 0.95 |
| *Herpes all (HSV, HZV, varicella-zoster)* | *Black* | 0.12 | 0.06 | <.01 | 0.11 | 0.11 | 0.84 |
| *Herpes all (HSV, HZV, varicella-zoster)* | *Education: less than high-school* | 0.27 | 0.22 | <.01 | 0.27 | 0.26 | 0.78 |
| *Herpes all (HSV, HZV, varicella-zoster)* | *Life insurance* | 0.64 | 0.68 | 0.06 | 0.65 | 0.66 | 0.53 |
| *Herpes all (HSV, HZV, varicella-zoster)* | *Smokes* | 0.09 | 0.06 | 0.01 | 0.09 | 0.09 | 0.93 |
| *Herpes all (HSV, HZV, varicella-zoster)* | *Excellent/good self-reported health* | 0.31 | 0.35 | 0.01 | 0.31 | 0.31 | 0.84 |
| *Herpes simplex (HSV)* | *Hypothyroidism* | 0.15 | 0.26 | <.01 | 0.15 | 0.18 | 0.36 |
| *Herpes simplex (HSV)* | *Rheumatoid arthritis/ collagen vascular diseases* | 0.07 | 0.13 | <.01 | 0.07 | 0.06 | 0.67 |
| *Herpes simplex (HSV)* | *Obesity* | 0.05 | 0.11 | <.01 | 0.05 | 0.06 | 0.70 |
| *Herpes simplex (HSV)* | *Depression* | 0.10 | 0.22 | <.01 | 0.10 | 0.09 | 0.78 |
| *Herpes simplex (HSV)* | *Education: less than high-school* | 0.27 | 0.15 | <.01 | 0.27 | 0.28 | 0.76 |
| *Herpes simplex (HSV)* | *Long-term care insurance* | 0.15 | 0.23 | <.01 | 0.15 | 0.15 | 0.83 |
| *Herpes vaccine ( shingles)* | *Cardiac arrhythmias* | 0.19 | 0.24 | 0.04 | 0.19 | 0.16 | 0.24 |
| *Herpes vaccine ( shingles)* | *Male* | 0.45 | 0.39 | 0.07 | 0.45 | 0.49 | 0.45 |
| *Herpes vaccine ( shingles)* | *Black* | 0.12 | 0.03 | <.01 | 0.11 | 0.15 | 0.53 |
| *Herpes vaccine ( shingles)* | *Education: some college* | 0.13 | 0.24 | <.01 | 0.14 | 0.12 | 0.33 |
| *Herpes vaccine ( shingles)* | *Lowest wealth quartile* | 0.22 | 0.09 | <.01 | 0.22 | 0.28 | 0.23 |
| *Herpes vaccine ( shingles)* | *Long-term care insurance* | 0.14 | 0.25 | <.01 | 0.15 | 0.13 | 0.48 |
| *Herpes zoster (HZV, shingles)* | *Hypothyroidism* | 0.14 | 0.22 | <.01 | 0.15 | 0.15 | 0.99 |
| *Herpes zoster (HZV, shingles)* | *Lymphoma* | 0.01 | 0.02 | 0.02 | 0.01 | 0.01 | 0.89 |
| *Herpes zoster (HZV, shingles)* | *Congestive heart failure* | 0.14 | 0.18 | <.01 | 0.14 | 0.14 | 0.87 |
| *Herpes zoster (HZV, shingles)* | *Rheumatoid arthritis/ collagen vascular diseases* | 0.06 | 0.11 | <.01 | 0.07 | 0.07 | 0.79 |
| *Herpes zoster (HZV, shingles)* | *Born before 1924* | 0.27 | 0.22 | 0.01 | 0.26 | 0.26 | 0.80 |
| *Herpes zoster (HZV, shingles)* | *Male* | 0.46 | 0.38 | <.01 | 0.45 | 0.45 | 0.95 |
| *Herpes zoster (HZV, shingles)* | *Black* | 0.12 | 0.07 | <.01 | 0.11 | 0.11 | 0.98 |
| *Herpes zoster (HZV, shingles)* | *Life insurance* | 0.64 | 0.69 | 0.04 | 0.65 | 0.66 | 0.64 |
| *Herpes zoster (HZV, shingles)* | *Smokes* | 0.09 | 0.06 | 0.01 | 0.09 | 0.09 | 0.85 |
| *Herpes zoster (HZV, shingles)* | *Excellent/good self-reported health* | 0.31 | 0.36 | 0.01 | 0.31 | 0.31 | 0.84 |
| *Pneumonia* | *Chronic pulmonary disease* | 0.17 | 0.48 | <.01 | 0.23 | 0.23 | 0.81 |
| *Pneumonia* | *Diabetes, uncomplicated* | 0.22 | 0.32 | <.01 | 0.24 | 0.24 | 0.75 |
| *Pneumonia* | *Hypothyroidism* | 0.14 | 0.20 | <.01 | 0.15 | 0.15 | 0.91 |
| *Pneumonia* | *Congestive heart failure* | 0.10 | 0.31 | <.01 | 0.14 | 0.14 | 0.82 |
| *Pneumonia* | *Solid tumor without metastasis* | 0.14 | 0.21 | <.01 | 0.15 | 0.16 | 0.82 |
| *Pneumonia* | *Rheumatoid arthritis/ collagen vascular diseases* | 0.06 | 0.10 | <.01 | 0.07 | 0.07 | 0.88 |
| *Pneumonia* | *Coagulopathy* | 0.04 | 0.10 | <.01 | 0.05 | 0.05 | 0.88 |
| *Pneumonia* | *Weight loss* | 0.04 | 0.08 | <.01 | 0.05 | 0.06 | 0.38 |
| *Pneumonia* | *Fluid and electrolyte disorders* | 0.10 | 0.25 | <.01 | 0.13 | 0.13 | 0.85 |
| *Pneumonia* | *Drug abuse* | 0.00 | 0.00 | 0.93 | 0.00 | 0.00 | 0.45 |
| *Pneumonia* | *Cardiac arrhythmias* | 0.16 | 0.28 | <.01 | 0.19 | 0.19 | 0.72 |
| *Pneumonia* | *Depression* | 0.09 | 0.15 | <.01 | 0.10 | 0.11 | 0.33 |
| *Pneumonia* | *Born 1924 - 1930* | 0.24 | 0.28 | 0.01 | 0.25 | 0.23 | 0.27 |
| *Pneumonia* | *Black* | 0.11 | 0.11 | 0.47 | 0.11 | 0.11 | 0.87 |
| *Pneumonia* | *Education: less than high-school* | 0.25 | 0.33 | <.01 | 0.27 | 0.26 | 0.51 |
| *Pneumonia* | *Education: graduate degree* | 0.07 | 0.08 | 0.95 | 0.07 | 0.07 | 0.65 |
| *Pneumonia* | *Heavy drinker* | 0.08 | 0.08 | 0.50 | 0.08 | 0.09 | 0.40 |
| *Pneumonia* | *Excellent/good self-reported health* | 0.27 | 0.47 | <.01 | 0.31 | 0.32 | 0.77 |
| *Pneumonia vaccine* | *Chronic pulmonary disease* | 0.21 | 0.28 | <.01 | 0.23 | 0.24 | 0.56 |
| *Pneumonia vaccine* | *Diabetes, uncomplicated* | 0.22 | 0.28 | <.01 | 0.24 | 0.25 | 0.56 |
| *Pneumonia vaccine* | *Hypothyroidism* | 0.13 | 0.19 | <.01 | 0.15 | 0.16 | 0.36 |
| *Pneumonia vaccine* | *Coagulopathy* | 0.04 | 0.07 | <.01 | 0.05 | 0.05 | 0.64 |
| *Pneumonia vaccine* | *Weight loss* | 0.04 | 0.07 | <.01 | 0.05 | 0.05 | 0.95 |
| *Pneumonia vaccine* | *Depression* | 0.09 | 0.13 | <.01 | 0.10 | 0.11 | 0.54 |
| *Pneumonia vaccine* | *Peripheral vascular disorders* | 0.14 | 0.19 | <.01 | 0.16 | 0.17 | 0.29 |
| *Pneumonia vaccine* | *Hypertension, uncomplicated* | 0.60 | 0.71 | <.01 | 0.63 | 0.64 | 0.62 |
| *Pneumonia vaccine* | *Born before 1924* | 0.34 | 0.09 | <.01 | 0.26 | 0.24 | 0.28 |
| *Pneumonia vaccine* | *Born 1924 - 1930* | 0.25 | 0.24 | 0.29 | 0.25 | 0.26 | 0.45 |
| *Pneumonia vaccine* | *Black* | 0.13 | 0.07 | <.01 | 0.11 | 0.10 | 0.30 |
| *Pneumonia vaccine* | *Hispanic* | 0.06 | 0.03 | <.01 | 0.05 | 0.06 | 0.56 |
| *Pneumonia vaccine* | *Married* | 0.61 | 0.69 | <.01 | 0.64 | 0.66 | 0.14 |
| *Pneumonia vaccine* | *Education: graduate degree* | 0.06 | 0.10 | <.01 | 0.08 | 0.08 | 0.72 |
| *Pneumonia vaccine* | *Veteran* | 0.34 | 0.29 | <.01 | 0.33 | 0.31 | 0.46 |
| *Pneumonia vaccine* | *Highest income quartile* | 0.24 | 0.26 | 0.04 | 0.25 | 0.26 | 0.38 |
| *Pneumonia vaccine* | *Lowest wealth quartile* | 0.24 | 0.16 | <.01 | 0.22 | 0.21 | 0.80 |
| *Pneumonia vaccine* | *Highest wealth quartile* | 0.24 | 0.32 | <.01 | 0.27 | 0.26 | 0.80 |
| *Pneumonia vaccine* | *Smokes* | 0.10 | 0.07 | <.01 | 0.09 | 0.09 | 0.67 |
| *Pneumonia vaccine* | *Three or more ADL limitations* | 0.05 | 0.04 | 0.02 | 0.05 | 0.05 | 0.43 |
| *Recurrent fungal infection (mycoses)* | *Diabetes, uncomplicated* | 0.21 | 0.39 | <.01 | 0.24 | 0.26 | 0.34 |
| *Recurrent fungal infection (mycoses)* | *Diabetes, complicated* | 0.06 | 0.23 | <.01 | 0.09 | 0.09 | 0.66 |
| *Recurrent fungal infection (mycoses)* | *Lymphoma* | 0.01 | 0.02 | <.01 | 0.01 | 0.01 | 0.95 |
| *Recurrent fungal infection (mycoses)* | *Rheumatoid arthritis/ collagen vascular diseases* | 0.06 | 0.11 | <.01 | 0.07 | 0.07 | 0.59 |
| *Recurrent fungal infection (mycoses)* | *Fluid and electrolyte disorders* | 0.11 | 0.22 | <.01 | 0.13 | 0.13 | 0.79 |
| *Recurrent fungal infection (mycoses)* | *Depression* | 0.09 | 0.17 | <.01 | 0.10 | 0.10 | 0.93 |
| *Recurrent fungal infection (mycoses)* | *Peripheral vascular disorders* | 0.13 | 0.32 | <.01 | 0.16 | 0.16 | 0.89 |
| *Recurrent fungal infection (mycoses)* | *Hypertension, uncomplicated* | 0.61 | 0.75 | <.01 | 0.63 | 0.64 | 0.67 |
| *Recurrent fungal infection (mycoses)* | *Male* | 0.46 | 0.38 | <.01 | 0.45 | 0.44 | 0.51 |
| *Recurrent fungal infection (mycoses)* | *Other race* | 0.02 | 0.01 | 0.03 | 0.02 | 0.02 | 0.78 |
| *Recurrent fungal infection (mycoses)* | *Education: less than high-school* | 0.27 | 0.27 | 0.82 | 0.27 | 0.27 | 0.81 |
| *Recurrent fungal infection (mycoses)* | *Education: some college* | 0.13 | 0.15 | 0.20 | 0.14 | 0.13 | 0.66 |
| *Recurrent fungal infection (mycoses)* | *Obese* | 0.22 | 0.32 | <.01 | 0.24 | 0.24 | 0.88 |
| *Recurrent fungal infection (mycoses)* | *One or two ADL limitations* | 0.11 | 0.19 | <.01 | 0.12 | 0.13 | 0.73 |
| *Recurrent fungal infection (mycoses)* | *Three or more ADL limitations* | 0.04 | 0.10 | <.01 | 0.05 | 0.05 | 0.97 |
| *Only those variables selected by the stepwise algorithm are shown. | | | | | | | |

| **Supplementary Table 5: Assessment of Pseudorandomization Quality - Alzheimer's Disease and Other Dementias*** | | | | | | | |
| --- | --- | --- | --- | --- | --- | --- | --- |
|  |  | **Before Pseudorandomization** | | | **After Pseudorandomization** | | |
|  |  | **Group** | | **χ2** | **Group** | | **χ2** |
| **Infection/Vaccine** | **Variable** | **Control** | **Treatment** | **p-val** | **Control** | **Treatment** | **p-val** |
| *Herpes all (HSV, HZV, varicella-zoster)* | *Diabetes, complicated* | 0.07 | 0.10 | <.01 | 0.07 | 0.08 | 0.90 |
| *Herpes all (HSV, HZV, varicella-zoster)* | *Hypothyroidism* | 0.13 | 0.23 | <.01 | 0.14 | 0.14 | 0.97 |
| *Herpes all (HSV, HZV, varicella-zoster)* | *Lymphoma* | 0.01 | 0.02 | <.01 | 0.01 | 0.01 | 0.89 |
| *Herpes all (HSV, HZV, varicella-zoster)* | *Rheumatoid arthritis/ collagen vascular diseases* | 0.06 | 0.11 | <.01 | 0.06 | 0.06 | 0.70 |
| *Herpes all (HSV, HZV, varicella-zoster)* | *Fluid and electrolyte disorders* | 0.11 | 0.16 | <.01 | 0.11 | 0.11 | 0.80 |
| *Herpes all (HSV, HZV, varicella-zoster)* | *Depression* | 0.08 | 0.14 | <.01 | 0.09 | 0.09 | 0.96 |
| *Herpes all (HSV, HZV, varicella-zoster)* | *Born before 1924* | 0.28 | 0.20 | <.01 | 0.27 | 0.26 | 0.73 |
| *Herpes all (HSV, HZV, varicella-zoster)* | *Male* | 0.46 | 0.37 | <.01 | 0.45 | 0.46 | 0.79 |
| *Herpes all (HSV, HZV, varicella-zoster)* | *Black* | 0.12 | 0.06 | <.01 | 0.11 | 0.11 | 0.87 |
| *Herpes all (HSV, HZV, varicella-zoster)* | *Education: less than high-school* | 0.27 | 0.21 | <.01 | 0.26 | 0.25 | 0.64 |
| *Herpes all (HSV, HZV, varicella-zoster)* | *Life insurance* | 0.64 | 0.68 | 0.05 | 0.65 | 0.66 | 0.56 |
| *Herpes all (HSV, HZV, varicella-zoster)* | *Smokes* | 0.09 | 0.06 | 0.01 | 0.09 | 0.09 | 0.84 |
| *Herpes all (HSV, HZV, varicella-zoster)* | *Excellent/good self-reported health* | 0.29 | 0.33 | 0.01 | 0.29 | 0.28 | 0.54 |
| *Herpes simplex (HSV)* | *Diabetes, complicated* | 0.07 | 0.13 | 0.01 | 0.07 | 0.08 | 0.68 |
| *Herpes simplex (HSV)* | *Hypothyroidism* | 0.14 | 0.26 | <.01 | 0.14 | 0.17 | 0.36 |
| *Herpes simplex (HSV)* | *Rheumatoid arthritis/ collagen vascular diseases* | 0.06 | 0.12 | 0.01 | 0.06 | 0.05 | 0.48 |
| *Herpes simplex (HSV)* | *Obesity* | 0.04 | 0.11 | <.01 | 0.05 | 0.06 | 0.41 |
| *Herpes simplex (HSV)* | *Depression* | 0.08 | 0.21 | <.01 | 0.09 | 0.09 | 0.76 |
| *Herpes simplex (HSV)* | *Born before 1924* | 0.27 | 0.16 | <.01 | 0.27 | 0.24 | 0.59 |
| *Herpes simplex (HSV)* | *Education: less than high-school* | 0.27 | 0.14 | <.01 | 0.26 | 0.24 | 0.67 |
| *Herpes simplex (HSV)* | *Long-term care insurance* | 0.15 | 0.23 | 0.01 | 0.15 | 0.15 | 0.94 |
| *Herpes vaccine ( shingles)* | *Cardiac arrhythmias* | 0.17 | 0.24 | 0.01 | 0.18 | 0.15 | 0.43 |
| *Herpes vaccine ( shingles)* | *Male* | 0.45 | 0.39 | 0.07 | 0.45 | 0.47 | 0.74 |
| *Herpes vaccine ( shingles)* | *Black* | 0.11 | 0.02 | <.01 | 0.11 | 0.14 | 0.61 |
| *Herpes vaccine ( shingles)* | *Education: some college* | 0.13 | 0.25 | <.01 | 0.14 | 0.12 | 0.38 |
| *Herpes vaccine ( shingles)* | *Education: graduate degree* | 0.07 | 0.12 | 0.03 | 0.08 | 0.07 | 0.87 |
| *Herpes vaccine ( shingles)* | *Lowest wealth quartile* | 0.21 | 0.09 | <.01 | 0.20 | 0.27 | 0.23 |
| *Herpes vaccine ( shingles)* | *Long-term care insurance* | 0.15 | 0.26 | <.01 | 0.15 | 0.14 | 0.63 |
| *Herpes zoster (HZV, shingles)* | *Hypothyroidism* | 0.14 | 0.22 | <.01 | 0.14 | 0.14 | 0.92 |
| *Herpes zoster (HZV, shingles)* | *Peptic ulcer disease excluding bleeding* | 0.03 | 0.05 | 0.02 | 0.03 | 0.04 | 0.76 |
| *Herpes zoster (HZV, shingles)* | *Rheumatoid arthritis/ collagen vascular diseases* | 0.06 | 0.11 | <.01 | 0.06 | 0.06 | 0.72 |
| *Herpes zoster (HZV, shingles)* | *Fluid and electrolyte disorders* | 0.11 | 0.17 | <.01 | 0.11 | 0.11 | 0.76 |
| *Herpes zoster (HZV, shingles)* | *Born before 1924* | 0.27 | 0.21 | <.01 | 0.27 | 0.26 | 0.75 |
| *Herpes zoster (HZV, shingles)* | *Male* | 0.46 | 0.37 | <.01 | 0.45 | 0.45 | 0.99 |
| *Herpes zoster (HZV, shingles)* | *Black* | 0.11 | 0.06 | <.01 | 0.11 | 0.11 | 0.99 |
| *Herpes zoster (HZV, shingles)* | *Life insurance* | 0.64 | 0.69 | 0.04 | 0.65 | 0.66 | 0.57 |
| *Herpes zoster (HZV, shingles)* | *Smokes* | 0.09 | 0.06 | 0.01 | 0.09 | 0.09 | 0.92 |
| *Herpes zoster (HZV, shingles)* | *Excellent/good self-reported health* | 0.29 | 0.34 | 0.02 | 0.29 | 0.29 | 0.84 |
| *Pneumonia* | *Chronic pulmonary disease* | 0.16 | 0.46 | <.01 | 0.22 | 0.22 | 0.84 |
| *Pneumonia* | *Diabetes, uncomplicated* | 0.21 | 0.30 | <.01 | 0.23 | 0.23 | 0.86 |
| *Pneumonia* | *Lymphoma* | 0.01 | 0.02 | <.01 | 0.01 | 0.01 | 0.95 |
| *Pneumonia* | *Congestive heart failure* | 0.09 | 0.28 | <.01 | 0.13 | 0.13 | 0.79 |
| *Pneumonia* | *Solid tumor without metastasis* | 0.13 | 0.21 | <.01 | 0.15 | 0.15 | 0.72 |
| *Pneumonia* | *Coagulopathy* | 0.04 | 0.10 | <.01 | 0.05 | 0.05 | 0.98 |
| *Pneumonia* | *Weight loss* | 0.04 | 0.07 | <.01 | 0.04 | 0.05 | 0.50 |
| *Pneumonia* | *Fluid and electrolyte disorders* | 0.09 | 0.22 | <.01 | 0.12 | 0.12 | 0.97 |
| *Pneumonia* | *Cardiac arrhythmias* | 0.15 | 0.27 | <.01 | 0.18 | 0.18 | 0.62 |
| *Pneumonia* | *Paralysis* | 0.03 | 0.07 | <.01 | 0.04 | 0.04 | 0.96 |
| *Pneumonia* | *Born 1924 - 1930* | 0.24 | 0.28 | 0.01 | 0.25 | 0.23 | 0.26 |
| *Pneumonia* | *Education: graduate degree* | 0.07 | 0.08 | 0.41 | 0.07 | 0.07 | 0.64 |
| *Pneumonia* | *Excellent/good self-reported health* | 0.25 | 0.44 | <.01 | 0.29 | 0.30 | 0.83 |
| *Pneumonia vaccine* | *Chronic pulmonary disease* | 0.19 | 0.27 | <.01 | 0.22 | 0.23 | 0.45 |
| *Pneumonia vaccine* | *Diabetes, uncomplicated* | 0.21 | 0.26 | <.01 | 0.23 | 0.24 | 0.45 |
| *Pneumonia vaccine* | *Hypothyroidism* | 0.13 | 0.19 | <.01 | 0.15 | 0.16 | 0.39 |
| *Pneumonia vaccine* | *Coagulopathy* | 0.04 | 0.06 | <.01 | 0.05 | 0.05 | 0.63 |
| *Pneumonia vaccine* | *Depression* | 0.08 | 0.12 | <.01 | 0.09 | 0.09 | 0.62 |
| *Pneumonia vaccine* | *Hypertension, uncomplicated* | 0.59 | 0.71 | <.01 | 0.62 | 0.63 | 0.63 |
| *Pneumonia vaccine* | *Born before 1924* | 0.34 | 0.09 | <.01 | 0.27 | 0.25 | 0.44 |
| *Pneumonia vaccine* | *Born 1924 - 1930* | 0.25 | 0.25 | 0.54 | 0.25 | 0.26 | 0.58 |
| *Pneumonia vaccine* | *Born 1942 - 1959* | 0.01 | 0.00 | 0.15 | 0.01 | 0.01 | 0.96 |
| *Pneumonia vaccine* | *Black* | 0.13 | 0.07 | <.01 | 0.11 | 0.10 | 0.41 |
| *Pneumonia vaccine* | *Hispanic* | 0.06 | 0.03 | <.01 | 0.05 | 0.06 | 0.50 |
| *Pneumonia vaccine* | *Married* | 0.62 | 0.70 | <.01 | 0.65 | 0.67 | 0.12 |
| *Pneumonia vaccine* | *Education: graduate degree* | 0.07 | 0.10 | <.01 | 0.08 | 0.08 | 0.84 |
| *Pneumonia vaccine* | *Veteran* | 0.34 | 0.30 | <.01 | 0.33 | 0.32 | 0.52 |
| *Pneumonia vaccine* | *Lowest wealth quartile* | 0.23 | 0.15 | <.01 | 0.20 | 0.20 | 1.00 |
| *Pneumonia vaccine* | *Highest wealth quartile* | 0.25 | 0.33 | <.01 | 0.27 | 0.27 | 0.70 |
| *Pneumonia vaccine* | *Long-term care insurance* | 0.14 | 0.19 | <.01 | 0.15 | 0.16 | 0.55 |
| *Pneumonia vaccine* | *Three or more ADL limitations* | 0.04 | 0.03 | 0.01 | 0.04 | 0.04 | 0.56 |
| *Recurrent fungal infection (mycoses)* | *Diabetes, uncomplicated* | 0.20 | 0.37 | <.01 | 0.23 | 0.25 | 0.31 |
| *Recurrent fungal infection (mycoses)* | *Diabetes, complicated* | 0.05 | 0.21 | <.01 | 0.08 | 0.08 | 0.58 |
| *Recurrent fungal infection (mycoses)* | *Lymphoma* | 0.01 | 0.02 | <.01 | 0.01 | 0.01 | 0.92 |
| *Recurrent fungal infection (mycoses)* | *Rheumatoid arthritis/ collagen vascular diseases* | 0.06 | 0.11 | <.01 | 0.07 | 0.07 | 0.60 |
| *Recurrent fungal infection (mycoses)* | *Fluid and electrolyte disorders* | 0.10 | 0.18 | <.01 | 0.12 | 0.12 | 0.76 |
| *Recurrent fungal infection (mycoses)* | *Depression* | 0.08 | 0.14 | <.01 | 0.09 | 0.09 | 0.96 |
| *Recurrent fungal infection (mycoses)* | *Peripheral vascular disorders* | 0.12 | 0.30 | <.01 | 0.14 | 0.15 | 0.83 |
| *Recurrent fungal infection (mycoses)* | *Hypertension, uncomplicated* | 0.60 | 0.74 | <.01 | 0.62 | 0.63 | 0.74 |
| *Recurrent fungal infection (mycoses)* | *Male* | 0.46 | 0.37 | <.01 | 0.45 | 0.44 | 0.63 |
| *Recurrent fungal infection (mycoses)* | *Education: less than high-school* | 0.26 | 0.26 | 0.77 | 0.26 | 0.26 | 0.75 |
| *Recurrent fungal infection (mycoses)* | *Education: some college* | 0.14 | 0.16 | 0.15 | 0.14 | 0.13 | 0.64 |
| *Recurrent fungal infection (mycoses)* | *Highest income quartile* | 0.25 | 0.26 | 0.74 | 0.25 | 0.26 | 0.68 |
| *Recurrent fungal infection (mycoses)* | *Obese* | 0.22 | 0.31 | <.01 | 0.24 | 0.24 | 0.78 |
| *Recurrent fungal infection (mycoses)* | *Poor/bad self-reported health* | 0.40 | 0.27 | <.01 | 0.38 | 0.37 | 0.73 |
| *Recurrent fungal infection (mycoses)* | *One or two ADL limitations* | 0.11 | 0.17 | <.01 | 0.12 | 0.13 | 0.52 |
| *Recurrent fungal infection (mycoses)* | *Three or more ADL limitations* | 0.03 | 0.07 | <.01 | 0.04 | 0.04 | 0.88 |
| *Only those variables selected by the stepwise algorithm are shown. | | | | | | | |

| **Supplementary Table 6: Assessment of Pseudorandomization Quality - Death*** | | | | | | | |
| --- | --- | --- | --- | --- | --- | --- | --- |
|  |  | **Before Pseudorandomization** | | | **After Pseudorandomization** | | |
|  |  | **Group** | | **χ2** | **Group** | | **χ2** |
| **Infection/Vaccine** | **Variable** | **Control** | **Treatment** | **p-val** | **Control** | **Treatment** | **p-val** |
| *Herpes all (HSV, HZV, varicella-zoster)* | *Diabetes, complicated* | 0.08 | 0.11 | <.01 | 0.09 | 0.09 | 0.46 |
| *Herpes all (HSV, HZV, varicella-zoster)* | *Hypothyroidism* | 0.14 | 0.23 | <.01 | 0.15 | 0.15 | 0.90 |
| *Herpes all (HSV, HZV, varicella-zoster)* | *Lymphoma* | 0.01 | 0.02 | <.01 | 0.01 | 0.01 | 0.90 |
| *Herpes all (HSV, HZV, varicella-zoster)* | *Solid tumor without metastasis* | 0.15 | 0.18 | 0.02 | 0.15 | 0.16 | 0.70 |
| *Herpes all (HSV, HZV, varicella-zoster)* | *Rheumatoid arthritis/ collagen vascular diseases* | 0.06 | 0.11 | <.01 | 0.07 | 0.07 | 0.90 |
| *Herpes all (HSV, HZV, varicella-zoster)* | *Depression* | 0.10 | 0.16 | <.01 | 0.11 | 0.11 | 0.70 |
| *Herpes all (HSV, HZV, varicella-zoster)* | *Born before 1924* | 0.27 | 0.21 | <.01 | 0.26 | 0.26 | 0.77 |
| *Herpes all (HSV, HZV, varicella-zoster)* | *Male* | 0.46 | 0.38 | <.01 | 0.45 | 0.45 | 0.97 |
| *Herpes all (HSV, HZV, varicella-zoster)* | *Black* | 0.12 | 0.07 | <.01 | 0.12 | 0.12 | 0.93 |
| *Herpes all (HSV, HZV, varicella-zoster)* | *Hispanic* | 0.06 | 0.04 | 0.13 | 0.05 | 0.05 | 0.95 |
| *Herpes all (HSV, HZV, varicella-zoster)* | *Life insurance* | 0.64 | 0.68 | 0.07 | 0.65 | 0.66 | 0.60 |
| *Herpes all (HSV, HZV, varicella-zoster)* | *Smokes* | 0.09 | 0.06 | 0.01 | 0.09 | 0.09 | 0.97 |
| *Herpes all (HSV, HZV, varicella-zoster)* | *Excellent/good self-reported health* | 0.31 | 0.36 | 0.02 | 0.32 | 0.32 | 0.86 |
| *Herpes simplex (HSV)* | *Hypothyroidism* | 0.15 | 0.26 | <.01 | 0.15 | 0.18 | 0.31 |
| *Herpes simplex (HSV)* | *Rheumatoid arthritis/ collagen vascular diseases* | 0.07 | 0.13 | <.01 | 0.07 | 0.06 | 0.60 |
| *Herpes simplex (HSV)* | *Obesity* | 0.05 | 0.11 | <.01 | 0.05 | 0.05 | 0.72 |
| *Herpes simplex (HSV)* | *Depression* | 0.10 | 0.23 | <.01 | 0.11 | 0.10 | 0.77 |
| *Herpes simplex (HSV)* | *Education: less than high-school* | 0.27 | 0.15 | <.01 | 0.27 | 0.29 | 0.71 |
| *Herpes simplex (HSV)* | *Long-term care insurance* | 0.14 | 0.23 | <.01 | 0.15 | 0.15 | 0.79 |
| *Herpes vaccine ( shingles)* | *Congestive heart failure* | 0.14 | 0.17 | 0.31 | 0.14 | 0.14 | 0.97 |
| *Herpes vaccine ( shingles)* | *Male* | 0.45 | 0.39 | 0.07 | 0.45 | 0.49 | 0.38 |
| *Herpes vaccine ( shingles)* | *Black* | 0.12 | 0.03 | <.01 | 0.12 | 0.14 | 0.61 |
| *Herpes vaccine ( shingles)* | *Education: some college* | 0.13 | 0.24 | <.01 | 0.13 | 0.12 | 0.52 |
| *Herpes vaccine ( shingles)* | *Education: graduate degree* | 0.07 | 0.11 | 0.02 | 0.07 | 0.08 | 0.97 |
| *Herpes vaccine ( shingles)* | *Lowest wealth quartile* | 0.23 | 0.10 | <.01 | 0.22 | 0.28 | 0.29 |
| *Herpes vaccine ( shingles)* | *Long-term care insurance* | 0.14 | 0.24 | <.01 | 0.15 | 0.13 | 0.54 |
| *Herpes zoster (HZV, shingles)* | *Hypothyroidism* | 0.15 | 0.22 | <.01 | 0.15 | 0.15 | 0.98 |
| *Herpes zoster (HZV, shingles)* | *Lymphoma* | 0.01 | 0.02 | 0.02 | 0.01 | 0.01 | 0.90 |
| *Herpes zoster (HZV, shingles)* | *Congestive heart failure* | 0.14 | 0.19 | <.01 | 0.14 | 0.15 | 0.70 |
| *Herpes zoster (HZV, shingles)* | *Rheumatoid arthritis/ collagen vascular diseases* | 0.06 | 0.11 | <.01 | 0.07 | 0.07 | 0.81 |
| *Herpes zoster (HZV, shingles)* | *Born before 1924* | 0.27 | 0.21 | 0.01 | 0.26 | 0.26 | 0.85 |
| *Herpes zoster (HZV, shingles)* | *Male* | 0.46 | 0.38 | <.01 | 0.45 | 0.45 | 0.93 |
| *Herpes zoster (HZV, shingles)* | *Black* | 0.12 | 0.07 | <.01 | 0.12 | 0.12 | 0.98 |
| *Herpes zoster (HZV, shingles)* | *Life insurance* | 0.64 | 0.68 | 0.05 | 0.65 | 0.65 | 0.72 |
| *Herpes zoster (HZV, shingles)* | *Smokes* | 0.09 | 0.06 | 0.01 | 0.09 | 0.09 | 0.93 |
| *Herpes zoster (HZV, shingles)* | *Excellent/good self-reported health* | 0.31 | 0.37 | 0.01 | 0.32 | 0.32 | 0.85 |
| *Pneumonia* | *Chronic pulmonary disease* | 0.17 | 0.48 | <.01 | 0.23 | 0.24 | 0.87 |
| *Pneumonia* | *Diabetes, uncomplicated* | 0.22 | 0.32 | <.01 | 0.24 | 0.25 | 0.78 |
| *Pneumonia* | *Congestive heart failure* | 0.10 | 0.32 | <.01 | 0.15 | 0.15 | 0.80 |
| *Pneumonia* | *Solid tumor without metastasis* | 0.14 | 0.21 | <.01 | 0.15 | 0.16 | 0.78 |
| *Pneumonia* | *Rheumatoid arthritis/ collagen vascular diseases* | 0.06 | 0.10 | <.01 | 0.07 | 0.07 | 0.77 |
| *Pneumonia* | *Coagulopathy* | 0.04 | 0.10 | <.01 | 0.05 | 0.05 | 0.97 |
| *Pneumonia* | *Weight loss* | 0.04 | 0.09 | <.01 | 0.05 | 0.06 | 0.44 |
| *Pneumonia* | *Fluid and electrolyte disorders* | 0.10 | 0.26 | <.01 | 0.14 | 0.14 | 0.86 |
| *Pneumonia* | *Cardiac arrhythmias* | 0.16 | 0.29 | <.01 | 0.19 | 0.19 | 0.72 |
| *Pneumonia* | *Psychoses* | 0.01 | 0.04 | <.01 | 0.02 | 0.02 | 0.93 |
| *Pneumonia* | *Born 1924 - 1930* | 0.24 | 0.28 | 0.01 | 0.25 | 0.23 | 0.34 |
| *Pneumonia* | *Black* | 0.12 | 0.11 | 0.57 | 0.12 | 0.12 | 0.89 |
| *Pneumonia* | *Education: less than high-school* | 0.25 | 0.34 | <.01 | 0.27 | 0.26 | 0.45 |
| *Pneumonia* | *Education: graduate degree* | 0.08 | 0.07 | 0.81 | 0.07 | 0.07 | 0.67 |
| *Pneumonia* | *Heavy drinker* | 0.08 | 0.08 | 0.59 | 0.08 | 0.09 | 0.34 |
| *Pneumonia* | *Excellent/good self-reported health* | 0.28 | 0.48 | <.01 | 0.32 | 0.33 | 0.75 |
| *Pneumonia vaccine* | *Chronic pulmonary disease* | 0.21 | 0.28 | <.01 | 0.23 | 0.24 | 0.71 |
| *Pneumonia vaccine* | *Diabetes, uncomplicated* | 0.22 | 0.28 | <.01 | 0.24 | 0.25 | 0.67 |
| *Pneumonia vaccine* | *Hypothyroidism* | 0.13 | 0.19 | <.01 | 0.15 | 0.16 | 0.41 |
| *Pneumonia vaccine* | *Metastatic cancer* | 0.02 | 0.03 | 0.02 | 0.02 | 0.02 | 0.95 |
| *Pneumonia vaccine* | *Coagulopathy* | 0.04 | 0.07 | <.01 | 0.05 | 0.05 | 0.70 |
| *Pneumonia vaccine* | *Weight loss* | 0.05 | 0.07 | <.01 | 0.05 | 0.05 | 0.84 |
| *Pneumonia vaccine* | *Depression* | 0.09 | 0.14 | <.01 | 0.11 | 0.11 | 0.66 |
| *Pneumonia vaccine* | *Peripheral vascular disorders* | 0.14 | 0.20 | <.01 | 0.16 | 0.17 | 0.39 |
| *Pneumonia vaccine* | *Hypertension, uncomplicated* | 0.60 | 0.71 | <.01 | 0.63 | 0.64 | 0.82 |
| *Pneumonia vaccine* | *Born before 1924* | 0.34 | 0.08 | <.01 | 0.26 | 0.25 | 0.39 |
| *Pneumonia vaccine* | *Born 1924 - 1930* | 0.25 | 0.24 | 0.41 | 0.25 | 0.26 | 0.48 |
| *Pneumonia vaccine* | *Black* | 0.13 | 0.07 | <.01 | 0.11 | 0.10 | 0.21 |
| *Pneumonia vaccine* | *Hispanic* | 0.06 | 0.04 | <.01 | 0.05 | 0.07 | 0.28 |
| *Pneumonia vaccine* | *Married* | 0.61 | 0.68 | <.01 | 0.63 | 0.65 | 0.25 |
| *Pneumonia vaccine* | *Education: graduate degree* | 0.06 | 0.10 | <.01 | 0.08 | 0.08 | 0.76 |
| *Pneumonia vaccine* | *Veteran* | 0.34 | 0.29 | <.01 | 0.32 | 0.31 | 0.34 |
| *Pneumonia vaccine* | *Highest income quartile* | 0.24 | 0.26 | 0.03 | 0.25 | 0.26 | 0.43 |
| *Pneumonia vaccine* | *Lowest wealth quartile* | 0.25 | 0.17 | <.01 | 0.22 | 0.22 | 0.97 |
| *Pneumonia vaccine* | *Highest wealth quartile* | 0.24 | 0.32 | <.01 | 0.26 | 0.26 | 0.74 |
| *Pneumonia vaccine* | *Smokes* | 0.10 | 0.07 | <.01 | 0.09 | 0.09 | 0.66 |
| *Pneumonia vaccine* | *Three or more ADL limitations* | 0.06 | 0.04 | 0.02 | 0.06 | 0.07 | 0.29 |
| *Recurrent fungal infection (mycoses)* | *Diabetes, uncomplicated* | 0.21 | 0.39 | <.01 | 0.24 | 0.26 | 0.27 |
| *Recurrent fungal infection (mycoses)* | *Diabetes, complicated* | 0.06 | 0.22 | <.01 | 0.09 | 0.09 | 0.64 |
| *Recurrent fungal infection (mycoses)* | *Lymphoma* | 0.01 | 0.02 | <.01 | 0.01 | 0.01 | 0.93 |
| *Recurrent fungal infection (mycoses)* | *Rheumatoid arthritis/ collagen vascular diseases* | 0.06 | 0.11 | <.01 | 0.07 | 0.08 | 0.59 |
| *Recurrent fungal infection (mycoses)* | *Fluid and electrolyte disorders* | 0.12 | 0.24 | <.01 | 0.14 | 0.14 | 0.72 |
| *Recurrent fungal infection (mycoses)* | *Depression* | 0.09 | 0.19 | <.01 | 0.11 | 0.11 | 0.94 |
| *Recurrent fungal infection (mycoses)* | *Peripheral vascular disorders* | 0.13 | 0.33 | <.01 | 0.16 | 0.16 | 0.99 |
| *Recurrent fungal infection (mycoses)* | *Hypertension, uncomplicated* | 0.61 | 0.73 | <.01 | 0.63 | 0.65 | 0.50 |
| *Recurrent fungal infection (mycoses)* | *Male* | 0.46 | 0.38 | <.01 | 0.45 | 0.44 | 0.50 |
| *Recurrent fungal infection (mycoses)* | *Hispanic* | 0.05 | 0.08 | <.01 | 0.06 | 0.06 | 0.98 |
| *Recurrent fungal infection (mycoses)* | *Education: less than high-school* | 0.27 | 0.27 | 0.96 | 0.27 | 0.27 | 0.91 |
| *Recurrent fungal infection (mycoses)* | *Education: some college* | 0.13 | 0.15 | 0.15 | 0.13 | 0.13 | 0.66 |
| *Recurrent fungal infection (mycoses)* | *Highest income quartile* | 0.24 | 0.24 | 0.64 | 0.24 | 0.25 | 0.88 |
| *Recurrent fungal infection (mycoses)* | *Obese* | 0.22 | 0.31 | <.01 | 0.24 | 0.24 | 0.88 |
| *Recurrent fungal infection (mycoses)* | *One or two ADL limitations* | 0.11 | 0.18 | <.01 | 0.12 | 0.13 | 0.62 |
| *Recurrent fungal infection (mycoses)* | *Three or more ADL limitations* | 0.04 | 0.12 | <.01 | 0.06 | 0.06 | 0.88 |
| *Only those variables selected by the stepwise algorithm are shown. | | | | | | | |
